# Breaking the 7 Mb barrier: Clinical cohort validation of genome-wide NIPT with fetal fraction enrichment and BinDel for detection of 1 Mb microdeletions and - duplications

**DOI:** 10.64898/2026.02.10.26345955

**Authors:** Karmen Vaiküll, Priit Paluoja, Signe Mölder, Vadym Gryshchenko, Neeme Tõnisson, Andres Salumets, Kaarel Krjutškov

## Abstract

**Objective:** To evaluate the analytical and clinical performance of fetal fraction (FF) enriched genome-wide noninvasive prenatal testing (GW-NIPT) for detection of clinically relevant copy number variants (CNVs) down to 1 Mb.

**Methods:** We retrospectively analyzed 10,501 singleton pregnancies tested with FF enrichment-based GW-NIPT between August 2023 and July 2025. CNV analysis was performed using BinDel and WisecondorX.

**Results:** FF enrichment increased median FF to 24% (2.4-fold increase). Clinically relevant CNVs, including microdeletions and microduplications, were reliably detected down to 1 Mb. Performance was robust across all maternal body mass index (BMI) categories. The retest rate was 0.95%, resulting in a final no-call rate of 0.03% with no BMI-attributable failures. The workflow demonstrated high sensitivity, specificity, and positive predictive value for common aneuploidies, rare autosomal trisomies, sex chromosome aneuploidies, subchromosomal CNVs, and pathogenic mitochondrial DNA variants.

**Conclusions:** FF enrichment enhances the analytical resolution of first-trimester GW-NIPT, enabling reliable detection of subchromosomal CNVs down to 1 Mb across diverse patient populations. This approach broadens the scope of prenatal screening while maintaining low test failure rates. All positive findings require confirmatory diagnostic testing and appropriate genetic counseling.

## Introduction

Non-invasive prenatal testing (NIPT) based on analysis of fetal-origin cell-free DNA (cfDNA) in maternal plasma has transformed prenatal screening for common aneuploidies such as trisomy 21 (T21), 18 (T18), and 13 (T13). Whole chromosomal abnormalities are detected with sensitivities and specificities exceeding 99%, establishing NIPT as a highly accurate screening assay for these conditions [1–3]. National implementation programmes, including the Dutch TRIDENT studies and the Belgian nationwide genome-wide (GW) NIPT programmes, have further demonstrated that GW-NIPT is feasible, robust, and clinically effective at the population scale, providing successfully karyotype-level information across all chromosomes [4,5]. However, Belgium’s national guidelines also highlight the complexity of genome-wide screening by recommending restrictive reporting of copy number variations (CNVs), limiting disclosure to clearly pathogenic findings with established clinical actionability [6]. These frameworks illustrate both the utility and interpretive challenges of GW-NIPT in routine clinical care.

A substantial proportion of clinically significant fetal chromosomal abnormalities arises from sub-chromosomal deletions and duplications, CNVs, rather than whole-chromosome aneuploidies. Sub-microscopic pathogenic CNVs, often under 10 Mb, are associated with congenital anomalies, intellectual disability, and neurodevelopmental disorders [7,8]. Although individually rare, their combined prevalence (∼1–2% of pregnancies) exceeds that of several common trisomies and contributes substantially to birth defects [9–12]. The majority of clinically actionable pathogenic CNVs identified in a population are smaller than 5 Mb, a size range that GW-NIPT often struggles to detect [13,14], raising the question of the optimal size threshold for CNV detection in NIPT.

Despite the strong performance of NIPT for whole chromosomal abnormalities, the reliable detection of CNVs <7 Mb remains technically challenging [15]. Small CNVs generate subtle deviations in sequencing read-depth that may be obscured by biological noise, maternal cfDNA predominance, naturally occurring random read distribution across the genome, and the limited resolution of low-coverage whole-genome sequencing [16,17]. Recent studies have identified ways for improvement: higher fetal fraction (FF) and optimized library preparation enhancing CNV detectability [7]; and advanced analytical pipelines capable of determining the fetal or maternal origin of detected aberrations significantly improving the positive predictive value for microdeletion syndromes [18,19]. Additionally, increasing sequencing depth has been shown to enhance sensitivity for detecting small CNVs [20]. Collectively, these findings underscore the need for methodological developments, such as FF enrichment and improved bioinformatic frameworks, to enable reliable detection of small, clinically relevant microdeletions and -duplications at higher genomic resolution.

BinDel is a targeted microdeletion risk detection tool focusing on clinically relevant pathogenic regions defined by carefully curated and validated genomic coordinates [19]. Using WG-NIPT data, BinDel models region-specific signal patterns and accounts for complex genomic architecture, including low-copy repeats and sequence homology. Validation studies demonstrated 88% sensitivity and 94% specificity [19]. In parallel, the non-targeted CNV detection algorithm WisecondorX has been adopted for NIPT to identify chromosomal imbalances [21]. The combined use of BinDel and WisecondorX therefore enables comprehensive CNV screening.

To date, pharmacogenetic objectives for newborns have not been incorporated into NIPT pipelines, despite their significant potential clinical value. For example, clinically actionable mitochondrial DNA (mtDNA) variants with direct implications for drug safety, including m.1555A>G variant associated with antibiotic, aminoglycoside-induced hearing loss are not screened in conventional NIPT, but have been included in some experimental NIPT studies [22,23]. FF enrichment increases the representation of mtDNA, which is typically more fragmented than genomic DNA [24,25]. This facilitates the assessment of these variants as an additional, clinically relevant analytical output with potential implications for the newborn, the mother, and additional offspring sharing maternal mitochondrial lineage.

We evaluated the analytical and clinical performance of a GW-NIPT approach incorporating FF enrichment to detect chromosomal abnormalities, including pathogenic CNVs as small as 1 Mb and the mitochondrial m.1555A>G variant. This study provides an integrated assessment of how FF enrichment improves analytical resolution and clinical utility in routine GW-NIPT.

## Methods

### Study design and cohort

In Estonia, NIPT has been part of the national prenatal screening programme since 2020, and the majority of NIPT samples generated nationally are analysed using the NIPTIFY test at the Celvia CC laboratory (Tartu, Estonia). The NIPTIFY assay is a GW-NIPT approach with proprietary laboratory procedures and data analysis pipelines [1,16,19,26,27].

This study comprised 10,501 singleton pregnancies from gestational week 10 onwards that underwent NIPTIFY testing between August 2023 and July 2025. Pregnancies with a vanishing twin (VT) were included when the demise of the co-twin was identified at or before 10 weeks of gestation or when the exact timing could not be determined. Ongoing twin pregnancies were excluded because the NIPTIFY analytical workflow is evaluated for pregnancies with a single ongoing fetus. The study was approved by the Research Ethics Committee of the University of Tartu (permit no. 403/M-14).

A total of 28 samples were excluded from the dataset used for performance calculations as exceptional cases, including four non-standard sex chromosome anomalies (confirmed XXYY, XXXXY, isodicentric Y-chromosome karyotypes, and a maternal-origin X0 mosaicism), two cases with inconclusive sex chromosome aneuploidy determination, four cases in which repeat sampling was requested but no follow-up sample was received, three no-call samples, and fifteen VT pregnancies. As a result, 10,473 samples were included in the performance calculations. For a subset of cases across different categories of common trisomies (T21, T18, T13), rare autosomal trisomies (RAT), CNVs, standard sex chromosome aneuploidies (SCAs), and m.1555A>G variant cases, confirmation status remained unknown due to loss to follow-up, lack of clinical feedback, or absence of confirmatory testing. These cases were excluded from category-specific performance calculations, and only confirmed true-positive, true-negative, and false-positive results were used, as no false-negative cases were identified.

Multiple findings within a single sample were handled at the sample level. Five samples contained two CNV findings and two samples contained three CNV findings per sample, each counted as a single sample. Three samples contained findings from two different categories (T21 and RAT, n=2; RAT and CNV, n=1), which were counted in both relevant categories. mtDNA analysis for the m.1555A>G variant was performed in 6,129 samples, reflecting the introduction of this screening component during the study period, of which 6,116 yielded a reportable result. Reported findings were considered confirmed when supported by invasive diagnostic testing, post-termination fetal tissue analysis, or clinical outcome information, including ultrasound findings and pregnancy loss. mtDNA findings were confirmed by Sanger-or amplicon-based Illumina sequencing.

### Clinical performance calculations

The following formulas were applied to calculate performance parameters. Sensitivity (true positive rate, TPR) was calculated as TP/(TP + FN), specificity (true negative rate, TNR) as TN/(TN + FP), and positive predictive value (PPV) as TP/(TP + FP), where TP denotes true positives, TN true negatives, FP false positives, and FN false negatives.

### Plasma processing and cell-free DNA extraction

Peripheral blood samples from pregnant women at ≥10 weeks of gestation were collected into cell-free DNA BCT tubes (Streck), and plasma was separated by standard dual centrifugation within seven days of blood collection [1]. For cell-free DNA extraction, 2 mL of plasma was transferred to a 24-well deep-well plate (Thermo Scientific) using a KingFisher APEX instrument. Subsequently, 100 µL of 20% SDS and 30 µL of Proteinase K (Omega Bio-tek) were added, after which the plate was sealed, vortexed, and incubated at 60°C for 30 minutes. Following incubation, JSB binding buffer and Mag-Bind Particles CH (Omega Bio-tek) were added according to the manufacturer’s instructions to complete the extraction. Cell-free DNA was eluted in 65 µL of elution buffer (5 mM Tris, pH 8.5).

### Library preparation

To generate the 32-plex libraries, end repair and A-tailing were performed using a mixture containing T4 PNK, T4 DNA polymerase, and Taq DNA polymerase in their respective buffer (Thermo Scientific [28]). A total of 25 µL of purified cell-free DNA was combined with 7 µL of the enzyme–buffer mix, sealed, vortexed, briefly centrifuged, and incubated sequentially at 20°C, 37°C, and 70°C for 10 minutes each (30 minutes total). Adapter ligation was carried out by adding 8 µL of T4 DNA ligase with ATP (Thermo Scientific) and 2 µL of 2.5 µM Illumina TruSeq forked indexed adapter (Metabion [29]), followed by incubation at 20°C for 1 hour. The ligated products were purified using an equal volume of Mag-Bind TotalPure NGS magnetic beads (Omega Bio-tek) with a 5-minute room-temperature incubation, and DNA was eluted in 20 µL of Milli-Q water. Subsequently, 8 µL of Blend Master Mix (Solis BioDyne) and 2 µL of a 10 µM Illumina P5/P7 primer mix (Metabion) were added. PCR was performed with an initial denaturation at 95°C for 12 minutes, followed by 12 cycles of 95°C for 20 seconds, 56°C for 20 seconds, and 72°C for 30 seconds, and a final extension at 72°C for 1 minute.

### Library quantification, size selection and sequencing

The concentration of each PCR product was measured using the Qubit BR assay (Thermo Scientific). PCR products were then pooled in equimolar amounts and size-selected on a 2% E-Gel SizeSelect II Agarose Gel (Thermo Scientific [25]) according to the manufacturer’s instructions. Electrophoresis was stopped when 250–260 bp fragments reached the pick well, after which the selected band was recovered and its size verified using the Agilent High Sensitivity D1000 ScreenTape assay. Libraries with fragment sizes between 250 and 260 bp were used for sequencing. It corresponds to 130 bp and 140 bp cfDNA size in **Figure 1**. Sequencing was performed on an Illumina NextSeq 1000 system using the NextSeq 1000/2000 P2 XLEAP-SBS reagent kit (100 cycles, Illumina), generating 80 bp single reads and six bp index reads.

**Figure 1.**
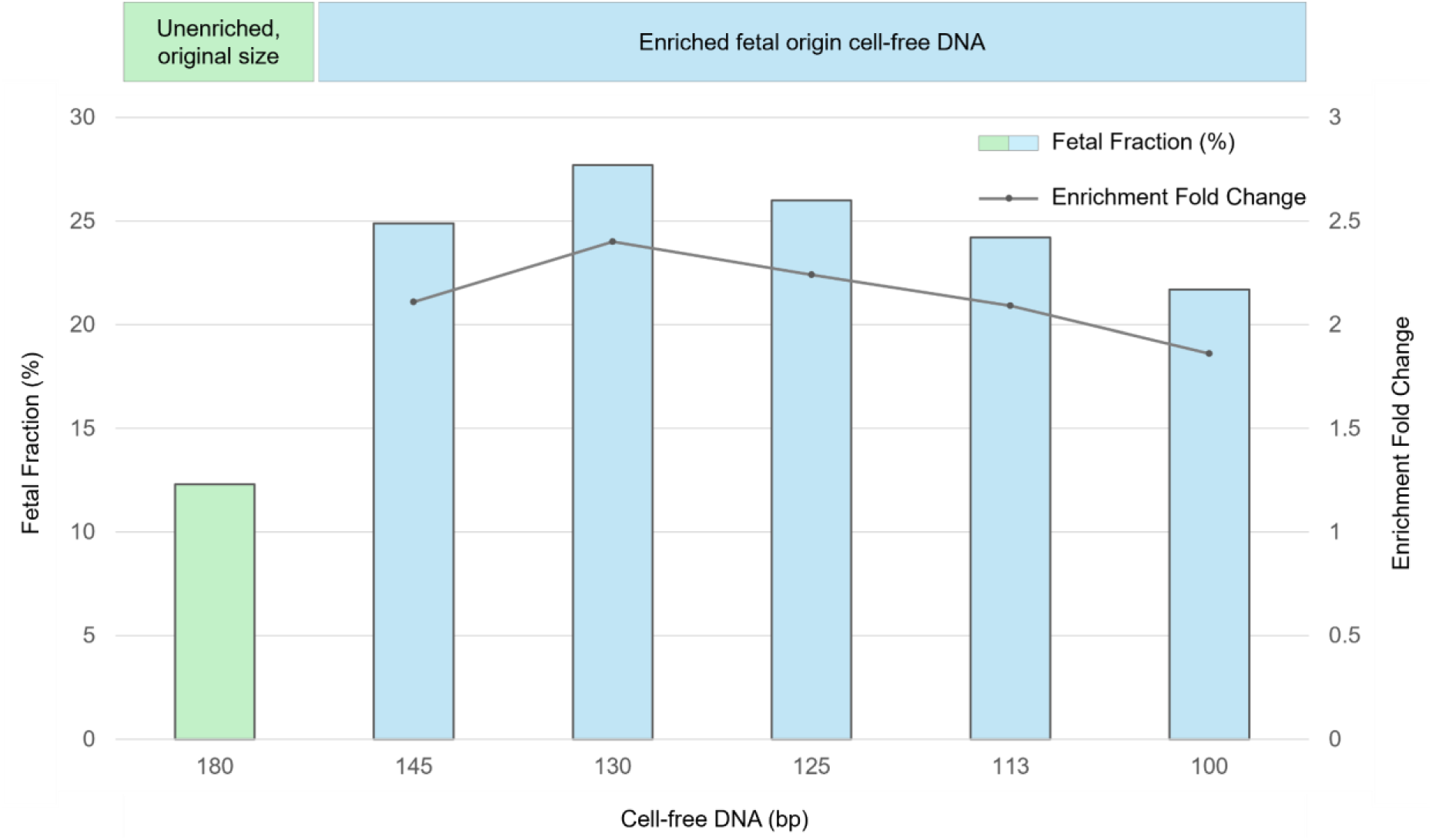
Relationship between cell-free DNA fragment length, fetal fraction, and enrichment efficiency. cfDNA fractions of different lengths are shown on the x-axis, Y-chromosome–based fetal fraction on the y-axis, and enrichment fold-change relative to the 180 bp unenriched cfDNA fraction on the z-axis.

### Computational analysis

Sequencing reads were processed through a bioinformatics pipeline managed using Nextflow 23.09.3-edge [30]. Briefly, reads were aligned to the human reference genome (GRCh38) using BWA-MEM 0.7.17 [31], and duplicate reads originating from a single fragment of DNA were marked and removed using GATK 4.2.5.0 MarkDuplicatesSpark [32]. Quality control required a minimum of six million aligned reads per sample [16]. Samples failing this threshold were resampled or resequenced. Chromosomal aneuploidies and sub-chromosomal CNVs were screened using BinDel (4.0.0) [19], NIPTeR (1.0.2) [33], NIPTmer [26], and WisecondorX (1.2.4) [21] softwares. FF was estimated using sex-specific methods as FFY for male fetuses based on Y-chromosome read quantification [34], and PREFACE (v0.1.2) [35] for female fetuses using genome-wide read distribution patterns. FF distributions and their association with maternal body mass index (BMI) were assessed using FFY only to maintain a homogeneous FF metric across samples. PREFACE-derived values for female fetuses were excluded to avoid mixing directly measured and model-derived estimates within the same analysis. Samples with FF <4% were flagged for repeat testing or further study. Mitochondrial m.1555A>G pharmacogenetic variant calling is based on pysam 0.22.1 [36] as described before [23].

## Results

### Fetal fraction enrichment

To assess the association between cfDNA fragment length and FF enrichment efficiency, we sequenced 13 maternal plasma samples from male-fetus pregnancies in technical duplicate. For each sample, one unenriched library (180 bp) and five size-enriched libraries (145, 130, 125, 113 and 100 bp) were prepared under identical conditions (**Figure 1**). The unenriched libraries showed a mean fetal fraction of 12.3%, whereas size enrichment increased FF, peaking at 130 bp (27.7%) and 125 bp (26.0%). Lower values of FF were observed at 145 bp (24.9%) and 113 bp (24.2%), with the lowest enrichment at 100 bp (21.7%). Enrichment fold-effects were highest at 130 bp (2.4) and 125 bp (2.2), and lowest at 100 bp (1.9). Based on these results, we selected the 130–140 bp size range for cfDNA enrichment for the clinical utility.

Sequencing coverage at the mitochondrial m.1555A>G position was evaluated in 13 maternal plasma samples using libraries constructed from cfDNA fragments of different lengths. In unenriched libraries, the mean coverage was 3× and increased progressively as cfDNA fragment length decreased, reaching a maximum mean coverage of 28× in libraries selected to 100 bp. In the clinical cohort assessed for the m.1555A>G variant (n = 6,129), NIPT performed with size selection resulted in a mean sequencing coverage of 21× at this position. A reportable result, defined as a minimum coverage of 1×, was obtained in 99.8% of cases (6,116/6,129).

Following FF selection of cell-free DNA, the mean Y-chromosome–derived fetal fraction among male fetuses in our clinical cohort was 23.9% (central 95% reference interval: 10.7%–39.0%) (**Figure 2A**). This represents a ∼2.6-fold increase compared with FFs reported in large European population-based datasets (**Supplementary Table S1**).

**Figure 2.**
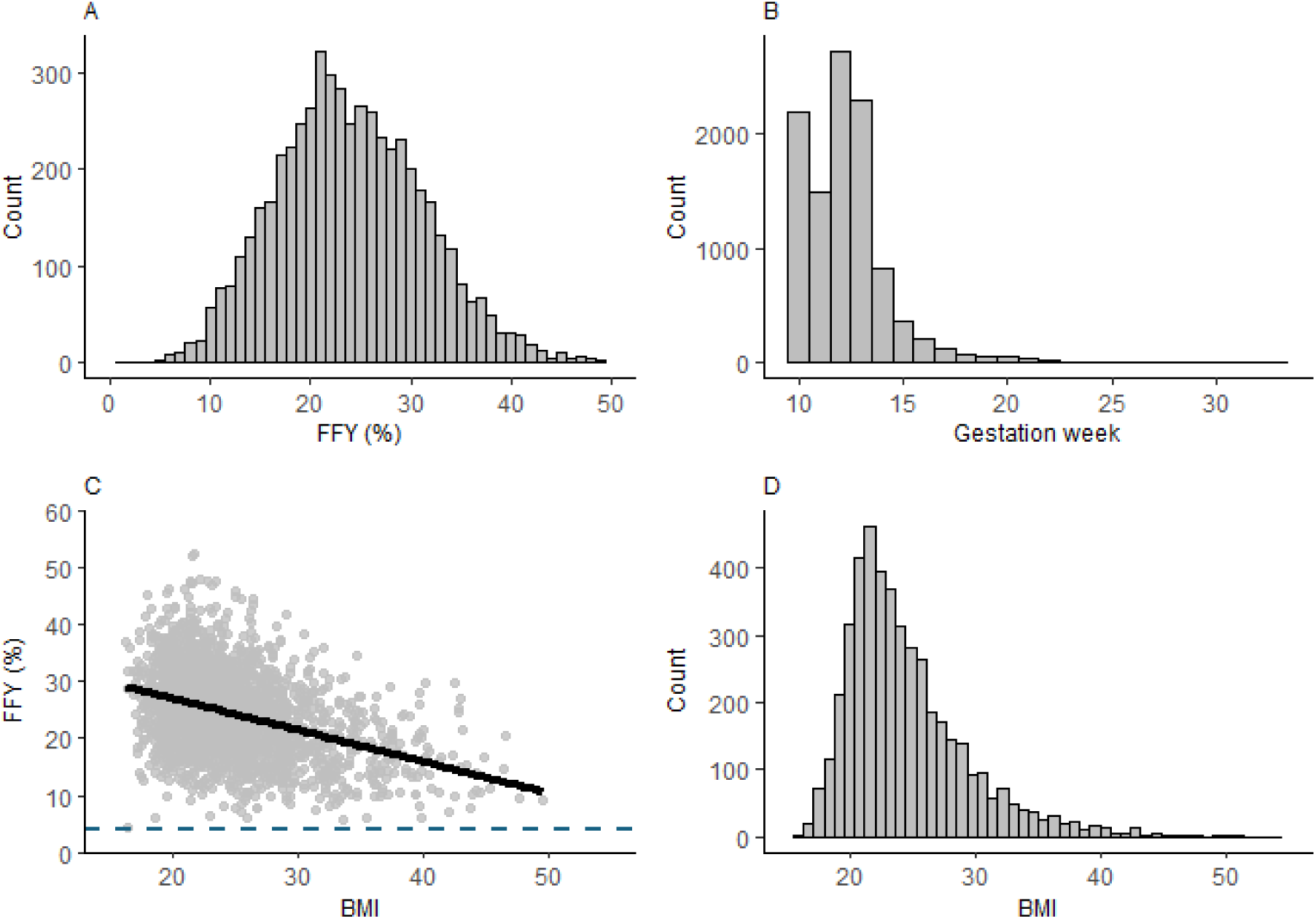
Characteristics of the study cohort. (A) Distribution of Y-chromosome-derived fetal fraction (FFY) in male fetuses. (B) Gestational age at sampling. (C) Association between enriched fetal fraction (FFY) and maternal body mass index (BMI); the solid line indicates the fitted trend. The dashed line shows a 4% fetal fraction threshold. (D) Distribution of maternal BMI in the study cohort.

### Clinical performance

The mean gestational age at the time of NIPT was 12.3 weeks (central 95% reference interval: 10–17 weeks) (**Figure 2B)**. To assess clinical test performance, a total of 10,473 samples were analysed. High-risk findings were reported in 244 cases (2.3%), and five cases were positive for the m.1555A>G variant. The confirmation information was unavailable for 75 high-risk cases (30.7%) and for two m.1555A>G variant cases. These were excluded from the performance-calculation dataset within their respective categories.

#### Common trisomies

A total of 68 T21 cases were identified, of which 50 were confirmed (**Table 1**), yielding a sensitivity of 100% (50/50). Two false-positive results were observed, while confirmatory data were unavailable for 16 cases. The specificity for T21 was 99.9% (10,405/10,407). Median z-scores (**Figure 3**) were consistent with the average FF of 23.9%.

**Figure 3.**
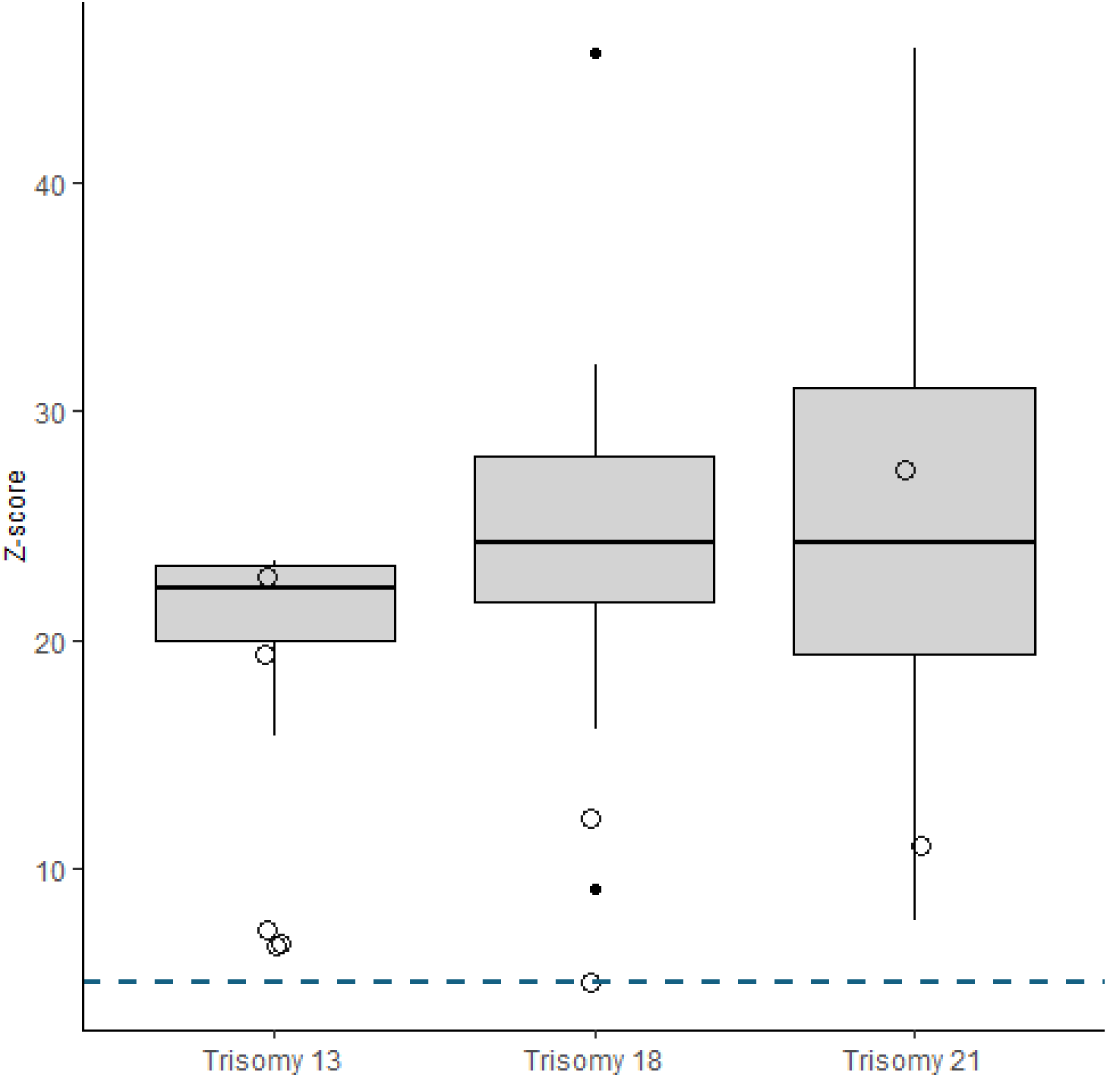
Z-score profiles of trisomy 13, 18 and 21 results. Boxplots represent the distribution of z-scores for true-positive cases across conditions. Open circles denote individual false-positive observations, overlaid with slight horizontal jitter to improve visibility. The horizontal dashed line indicates the z-score threshold of 5 used for classification.

**Table 1.** Sensitivity, specificity, and positive predictive value of the NIPT across three trisomy categories.

| Aberration | NA | TP | FP | TN | FN | TPR<br>(Sensitivity) | TNR<br>(Specificity) | PPV |
| --- | --- | --- | --- | --- | --- | --- | --- | --- |
| T21 | 16 | 50 | 2 | 10,405 | 0 | 100% | 99.98% | 96.15% |
| T18 | 5 | 16 | 2 | 10,450 | 0 | 100% | 99.98% | 88.89% |
| T13 | 3 | 4 | 5 | 10,461 | 0 | 100% | 99.95% | 44.44% |

*Abbreviations: NA, confirmation result not available; T21, trisomy 21; T18, trisomy 18; T13, trisomy 13; TP, true positive; FP, false positive; TN, true negative; FN, false negative; TPR, sensitivity; TNR, specificity; PPV, positive predictive value.*

Among the 23 T18 cases, 16 were confirmed, corresponding to a sensitivity of 100% (16/16). Two false-positive results were observed (**Figure 3)**, and five cases lacked confirmation information. Specificity for T18 was 99.9% (10,450/10,452).

Twelve T13 findings were reported, of which four were confirmed, demonstrating 100% sensitivity (4/4). Five false-positive results occurred (**Figure 3)**, and confirmation was unavailable for three cases. Specificity for T13 was 99.9% (10,461/10,466).

#### Copy-number variants

A total of 38 samples screened positive for CNVs. In 11 cases, findings were confirmed (**Table 2**), yielding a sensitivity of 100% (11/11) (**Table 3**). 12 samples were false positives, and 15 lacked confirmation. Reported CNVs included 26 deletions and 21 duplications ranging from 0.5 to 75.8 Mb in size, with confirmed CNVs ranging from 1.2 to 75.8 Mb (more than one abnormality were found in 7 cases). Specificity for CNVs was 99.9% (10,435/10,447).

**Table 2.** True-positive CNVs with gestational age, size, syndrome region involved and confirmation details. Samples 9–11 each contain two CNVs.

| Sample | Gest. age | FF | Fetal sex | CNV | Syndrome region | Size (Mb) | Confirmation |
| --- | --- | --- | --- | --- | --- | --- | --- |
| 1 | 10+3 | 21.98 | male | 17q11.2 del | NF1 microdeletion syndrome | 1.2 | AC |
| 2 | 13+5 | 25.82 | female | 17q12 del | 17q12 microdeletion syndrome | 1.4 | AC |
| 3 | 12+6 | 31.79 | male | 22q11.21 dup | 22q11.2 duplication syndrome | 2.1 | AC |
| 4 | 13+1 | 27.68 | female | 1p36.32-p36.31 del | 1p36 deletion syndrome | 3 | AC |
| 5 | 10+6 | 17.45 | female | 11q24.3-q25 del | partial Jacobsen syndrome | 3.2 | AC |
| 6 | 12+5 | 22.83 | female | 5p15.33-p15.31 del | Cri-Du-Chat syndrome | 8.4 | AC |
| 7 | 13+1 | 24.91 | female | 21q22.13-q22.3 dup | Partial trisomy 21 | 9.5 | AC |
| 8 | 13 | 17.04 | male | 9p24.3-p23 del | Distal monosomy 9p | 12.5 | AC |
| 9 | 11+4 | 29.73 | female | 5p15.33-p15.2 del | Cri-Du-Chat syndrome | 14.6 | CVS |
|  |  |  |  | 8p23.3-p21.2 dup | 8p23.1 duplication syndrome | 25.9 |  |
| 10 | 11+2 | 16.5 | female | 4p16.3-p15.1 del | Wolf-Hirschhorn syndrome | 31.1 | MC |
|  |  |  |  | 4q33-q35.2 del | Terminal 4q deletion | 20.1 |  |
| 11 | 11+5 | 25.46 | female | Xp22.33-p21.3 del |  | 28.8 | AC |
|  |  |  |  | Xq21.1-q28 dup |  | 75.8 |  |

*Abbreviations: CNV, copy number variant; FF, fetal fraction; AC, amniocentesis; CVS, chorionic villus sampling; MC, miscarriage; Gest. age, gestational age; del, deletion; dup, duplication.*

**Table 3.**
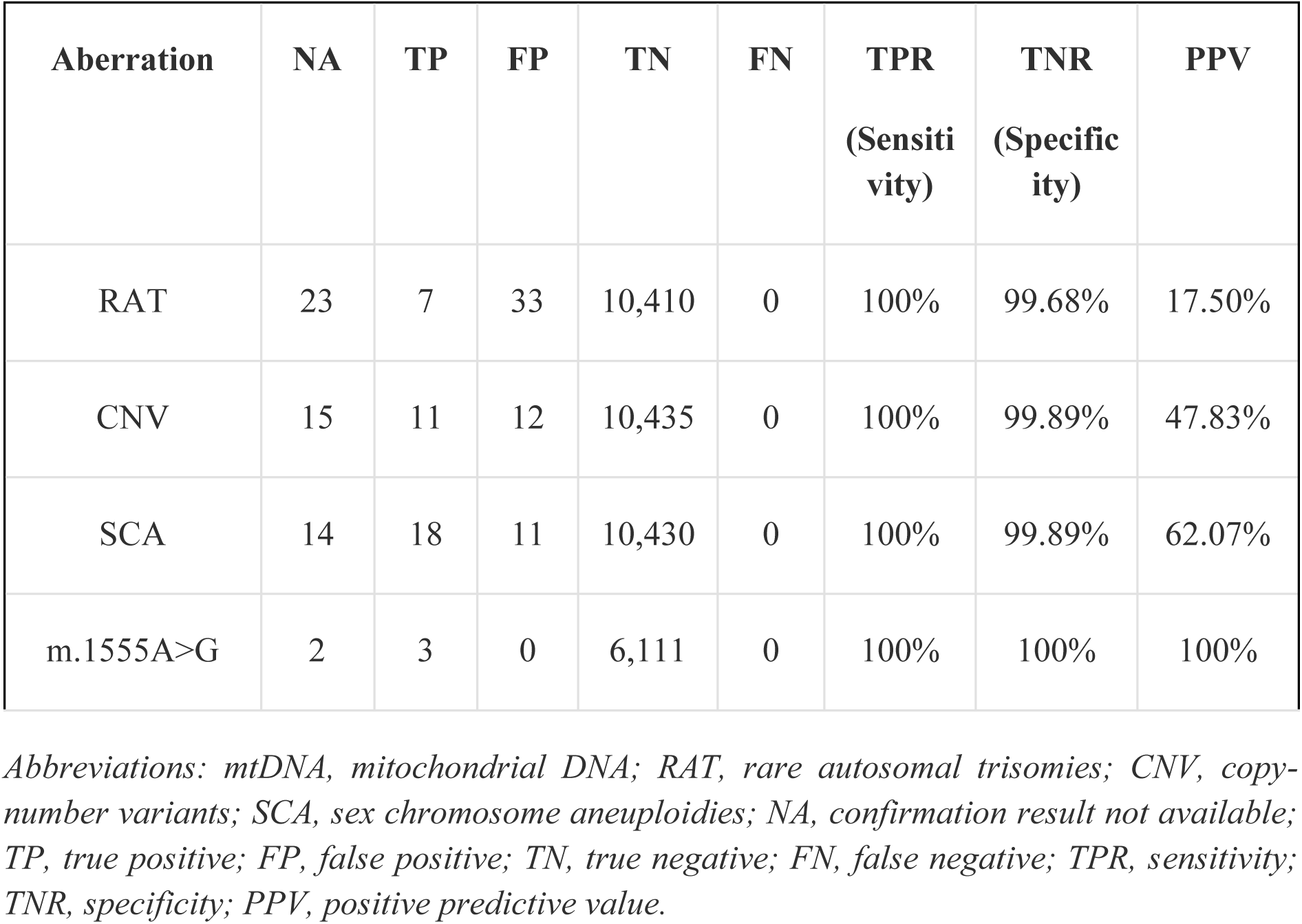
Sensitivity, specificity, and positive predictive value of the NIPT across major chromosomal abnormality categories and mtDNA point mutation.

#### Rare autosomal trisomies

Sixty-three samples screened positive for RATs (**Table 3**). Seven cases were confirmed (T22: n=4; T9: n=2; T16: n=1), yielding 100% sensitivity (7/7). There were 33 false-positive results, and confirmation was unavailable for 23 cases. Specificity for RAT detection was 99.7% (10,410/10,443).

#### Sex chromosome aneuploidies

Twenty-three samples were reported as monosomy X (X0), with eight confirmed, resulting in 100% sensitivity (8/8) (**Table 3**). Nine false positives were observed, and six cases had unknown confirmation status. Specificity was 99.9% (10,450/10,459).

Trisomy X (XXX) was identified in five samples; one case was confirmed, two were false positives, and two lacked confirmation status. Sensitivity was 100% (1/1) and specificity was 99.9% (10,468/10,470).

Eleven XXY cases were detected; 8 were confirmed, and three lacked confirmation status. Sensitivity was 100% (8/8), with no false positives, resulting in 100% specificity (10,462/10,462).

Four samples screened positive for XYY; one was confirmed, and three lacked confirmation status. Sensitivity was 100% (1/1), and specificity was 100% (10,469/10,469).

#### Mitochondrial DNA variant analysis

Mitochondrial analysis was available for 6,116 samples. The m.1555A>G variant was reported when the variant allele fraction exceeded 50%. Five samples screened positive; three underwent Sanger confirmation and were all verified as true positives; confirmatory data were unavailable for the remaining two cases. Sensitivity for mtDNA variant detection was 100% (3/3), and specificity was 100% (6,111/6,111).

### Vanishing twin

A total of 15 VT cases were identified in the study cohort (**Supplementary Table S2**). Chromosomal aneuploidy was detected in 33% of cases after the initial NIPT analysis (5/15; T13, n=2; T16, n=2; T15, n=1). Following repeat testing with samples collected at least two weeks later, of these five cases, the aneuploidy signal disappeared in three samples, indicating a possible association with demised twin. Two signals (T13 and T15) persisted and were reported. Follow-up evaluation revealed no fetal abnormalities in both cases.

Two additional VT cases showed low-level Y-chromosome signals. One was reported as low risk with undetermined fetal sex, while in the other, repeat sampling yielded a low Y signal; the pregnancy was subsequently confirmed by ultrasound to involve a surviving female fetus. Reportable results were obtained for all VT cases, with no no-call outcomes.

### No-call rates and BMI

Resampling was required in 100 cases (0.95%) due to inconclusive findings, reduced FF, elevated standard deviation (SD), VT, or low sequencing read count. After repeat testing, three samples remained non-reportable, resulting in a final no-call rate of 0.03% (3/10,501), all due to persistently elevated SD values above the predefined cutoff (SD >2).

The mean BMI in the study cohort was 24.4 (central 95% reference interval, 18.1–37.1; **Figure 2D**). After FF enrichment, FF values remained above the 4% cutoff across the BMI range despite the expected inverse association between BMI and FF (**Figure 2C**). No no-call outcomes were attributable to elevated BMI.

## Discussion

In this study, we demonstrate that FF enrichment combined with optimised software extends CNV detection resolution from the 7 Mb threshold of standard GW-NIPT [15,37,38] down to 1 Mb. By selecting cfDNA fragments in the 130–140 bp size range, we elevated the median FF to 24% in our cohort of 10,473 singleton pregnancies from gestational week 10 onward (**Figure 2A**). This enhanced resolution enabled detection of CNVs down to 1 Mb, addressing a clinically significant gap given that 79% of pathogenic CNVs are smaller than 5 Mb [13]. Additionally, enrichment of shorter cfDNA fragments increased mtDNA sequencing depth, enabling accurate genotyping of the pharmacogenetically relevant m.1555A>G variant.

The enhanced FF achieved through our methodology provides advantages for clinical applicability, particularly in challenging scenarios. High analytical sensitivity is essential for conditions such as T13 and T18, where z-scores are typically lower than those observed in T21 [39,40]. By maintaining consistently elevated fetal fraction regardless of maternal BMI (**Figure 2C**), our approach eliminated BMI-associated test failures, removing a known limitation of standard NIPT implementation [41]. Furthermore, our method demonstrated robustness in biologically complex pregnancies: all 15 VT cases in our cohort yielded reportable results with no no-call outcomes, despite transient aneuploidy signals in some cases (**Supplementary Table S2**). The final no-call rate following repeat sampling was 0.03%, substantially lower than the 0.3% reported in the TRIDENT-2 population-based study [4], highlighting the combined benefit of FF enrichment. Repeat sampling was requested in 0.95% of cases and served as a clinically appropriate strategy to resolve inconclusive results. Requesting a repeat blood draw following an initial non-reportable or ambiguous NIPT result serves the interests of both patients, clinicians, and the testing laboratory.

While the analytical sensitivity for all tested conditions reached a very high level in our cohort (**Table 1-3**), interpretation of positive predictive values requires careful consideration of biological and clinical context. The PPV for RATs was 17.5% (7/40 with known outcomes), consistent with the expected range from meta-analyses reporting pooled PPVs of approximately 11.5% [42]. This reflects the biological reality that most RAT signals represent confined placental mosaicism rather than true fetal aneuploidy. However, RAT detection retains clinical significance, as these findings are associated with increased risk of adverse pregnancy outcomes, including pre-eclampsia, preterm birth, and pregnancy loss, even if invasive testing does not prove the fetal involvement in aneuploidy [43], warranting appropriate counseling and monitoring. For CNVs, the observed PPV was 47.8% (11/23 with known outcomes), higher than meta-analytic estimates of 33–37% [44,45], although the limited sample size in this study precludes definitive conclusions. Nevertheless, these PPV findings underscore that GW NIPT remains a screening test requiring confirmatory diagnostic testing and multidisciplinary clinical judgment for positive results.

Implementation of GW-NIPT must be contextualized within broader healthcare frameworks that extend beyond laboratory analytical performance. National programs such as the Belgian NIPT guidelines and the Dutch TRIDENT-2 initiative represent system-level agreements governing reporting practices, post-test counseling, and decisions regarding invasive diagnostic procedures. These frameworks appropriately emphasize that laboratory findings must inform, rather than dictate, clinical management decisions. Evidence from the Netherlands indicates that most pregnant women opt for GW-NIPT when offered a choice [4], reflecting patient preference for comprehensive prenatal information. This preference supports the clinical rationale for broader screening panels when analytical performance is sufficient to maintain appropriate PPV thresholds.

A significant limitation of this study is incomplete clinical follow-up, which affected 30% of positive screening results. Post-NIPT feedback was limited by data protection regulations restricting information sharing from clinicians, by cases without confirmatory testing, and by patients who were lost to follow-up or declined further contact. This incomplete ascertainment creates potential interpretive gaps, as actual clinical outcomes cannot always be comprehensively verified. Consequently, the reported performance metrics, particularly specificity calculations, represent best-case estimates under the assumption that unconfirmed cases follow similar distributions to confirmed cases. This assumption may not hold if patients with false-positive results are more likely to decline follow-up than those with true-positive findings. Additionally, while no false negatives were identified among cases with known outcomes, the incomplete follow-up rate limits confidence in this finding, as missed diagnoses may be systematically underreported.

In conclusion, FF enrichment addresses key technical limitations of standard GW-NIPT by enabling the detection of 1 Mb CNVs, eliminating or minimizing BMI-related test failures, and maintaining performance in VT cases. These improvements expand the clinical utility of GW-NIPT, particularly in early gestational weeks and diverse patient populations. However, GW-NIPT remains a screening test in which biologically driven false-positive results are likely and false-negative results cannot be entirely excluded. Positive findings require confirmatory diagnostic testing, and implementation must be guided by multidisciplinary clinical frameworks that integrate analytical performance with patient counseling, informed consent, and appropriate follow-up protocols. The observed performance characteristics support GW-NIPT as a valuable screening tool when implemented within structured healthcare systems that prioritize comprehensive patient care over isolated laboratory metrics.

## Data Availability

The sequencing data used in this study are not publicly available due to concerns about data sensitivity. The data can be accessed from the Celvia CC Data Access Committee upon reasonable request, subject to approval from the Estonian Research Council committee, compliance with GDPR requirements, and permission from the Celvia CC Data Access Committee. The data are stored in controlled-access facilities.

## Acknowledgements

The authors express their sincere gratitude to the participating patients and health-care professionals involved in the study.

## Conflict of interest

The authors declare no competing interests.

## Funding statement

The present study was funded by the Estonian Research Council (grants no. PSG1082 and PRG1076); Horizon Europe grant (NESTOR, grant no. 101120075); Novo Nordisk Foundation (grant no. NNF24OC0092384), the Swedish Research Council (grant no. 2024-02530) and the Estonian Ministry of Education and Research Centres of Excellence grant TK214 name of CoE.

## Supplementary data

**Supplementary Table S1.**
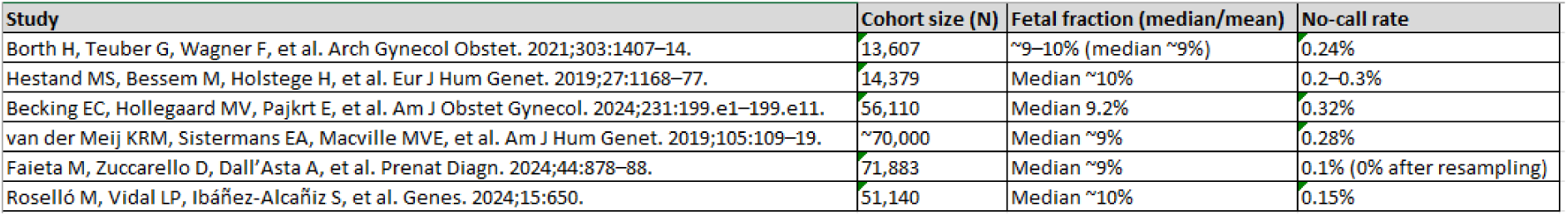
Summary of European population-based NIPT studies (>10,000 pregnancies) reporting fetal fraction and no-call rates.

| Study | Cohort size (N) | Fetal fraction (median/mean) | No-call rate |
| --- | --- | --- | --- |
| Borth H, Teuber G, Wagner F, et al. Arch Gynecol Obstet. 2021;303:1407–14. | 13,607 | ~9–10% (median ~9%) | 0.24% |
| Hestand MS, Bessem M, Holstege H, et al. Eur J Hum Genet. 2019;27:1168–77. | 14,379 | Median ~10% | 0.2–0.3% |
| Becking EC, Hollegaard MV, Pajkrt E, et al. Am J Obstet Gynecol. 2024;231:199.e1–199.e11. | 56,110 | Median 9.2% | 0.32% |
| van der Meij KRM, Sistermans EA, Macville MVE, et al. Am J Hum Genet. 2019;105:109–19. | ~70,000 | Median ~9% | 0.28% |
| Faieta M, Zuccarello D, Dall'Asta A, et al. Prenat Diagn. 2024;44:878–88. | 71,883 | Median ~9% | 0.1% (0% after resampling) |
| Roselló M, Vidal LP, Ibáñez-Alcañiz S, et al. Genes. 2024;15:650. | 51,140 | Median ~10% | 0.15% |

**Supplementary Table S2.**
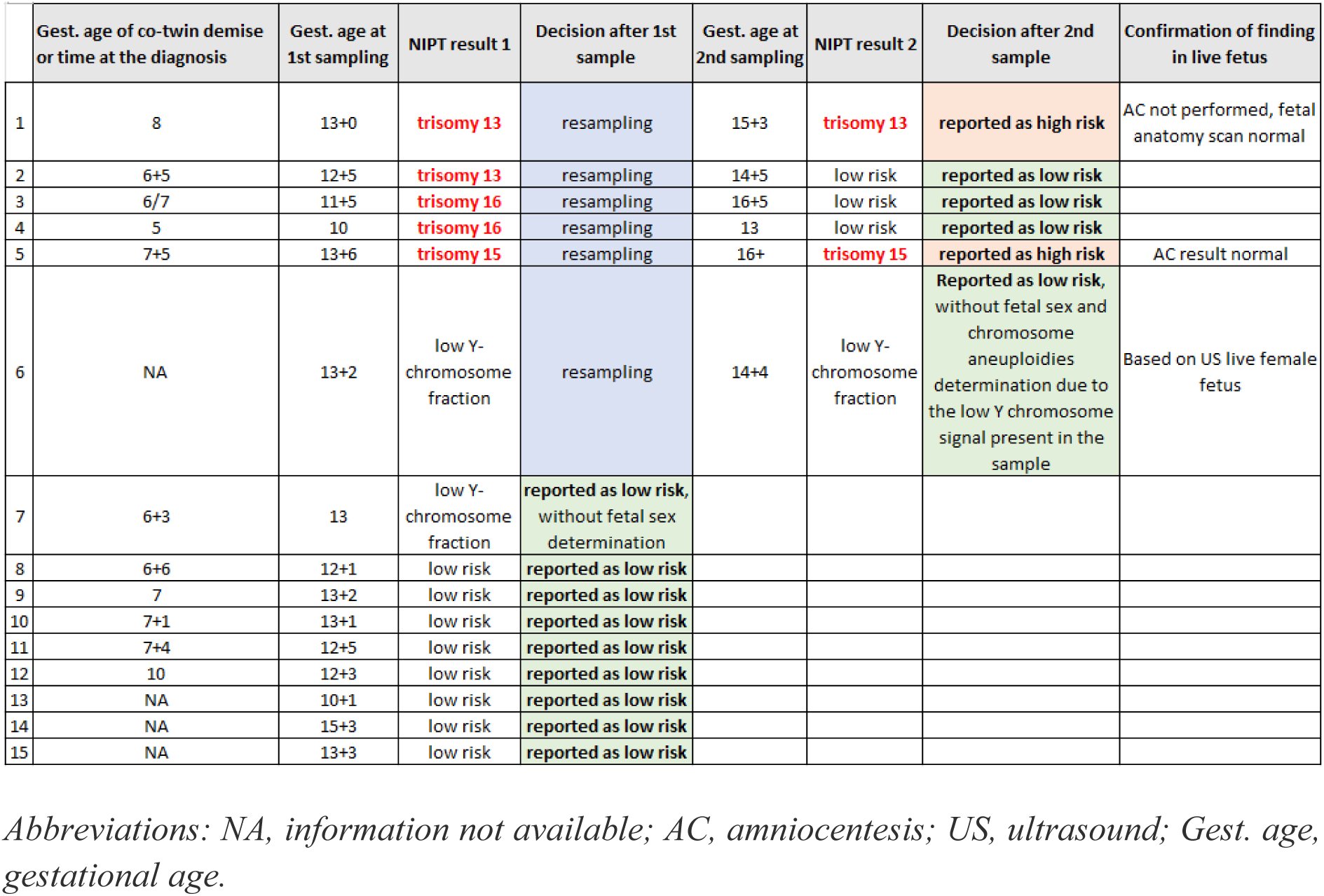
Summary of vanishing twin cases in the study cohort.

## Notes

### Competing Interest Statement

The authors have declared no competing interest.

### Author Declarations

The study was approved by the Research Ethics Committee of the University of Tartu (permit no. 403/M-14).

